# Reproduction as a Means of Evaluating Policy Models: A Case Study of a COVID-19 Simulation

**DOI:** 10.1101/2021.01.29.21250743

**Authors:** E. Chattoe-Brown, N. Gilbert, D. A. Robertson, C. Watts

**Affiliations:** School of Media, Communication and Sociology, University of Leicester; Department of Sociology, University of Surrey; School of Business and Economics, Loughborough University; St Catherine’s College, University of Oxford; Independent Researcher

**Keywords:** COVID-19, Epidemiological Models, Agent-Based Modelling, Methodology, Model Verification, Model Replication, Computational Social Science, Model Quality Assurance

## Abstract

This article proposes (and demonstrates the effectiveness of) a new strategy for assessing the results of epidemic models which we designate *reproduction*. The strategy is to build an independent model that uses (as far as possible) only the published information about the model to be assessed. In the example presented here, the independent model also follows a different modelling approach (agent-based modelling) to the model being assessed (the London School of Hygiene and Tropical Medicine compartmental model which has been influential in COVID lockdown policy). The argument runs that if the policy prescriptions of the two models match then this independently supports them (and reduces the chance that they are artefacts of assumptions, modelling approach or programming bugs). If, on the other hand, they do not match then either the model being assessed is not provided with sufficient information to be relied on or (perhaps) there is something wrong with it. In addition to justifying the approach, describing the two models and demonstrating the success of the approach, the article also discusses additional benefits of the reproduction strategy independent of whether match between policy prescriptions is actually achieved.

## 1. Introduction

The COVID-19 pandemic has given modelling a renewed policy relevance (Boland 2020, Currie *et al*. 2020, Ford 2020, Sample 2020). However, if models are to play a part in crucial decisions, such as the trade-off between deaths and the severe economic dislocation entailed by enforcing or relaxing “lock down” at particular times, then, so the argument runs, politicians and the public must be able to trust them.

The practicalities of modelling give rise to a major challenge for creating this trust. The models are large and complicated, with many assumptions and diverse sources of data. The models that serve as leading contenders to inform policy have usually evolved over many years.^1^ Once a pandemic is in progress, there may be inadequate time to peer review new model analyses, resulting in the *ad hoc* adaptation of models to meet new policy challenges (with an associated loss of transparency).

This article proposes that we need to step back and consider carefully how collective action can improve trust in a model. One way is to show that the same results can be obtained from a second, independently derived model based on shared behavioural assumptions. This is related conceptually to the traditional scientific strategy of replication (Edmonds and Hales 2003), in which the results of a study conducted by one team are checked against those obtained independently by another team. Distinctions can be made between different types of comparison. One might simply re-run a model simulation using the same code on different hardware. One might rewrite the code in a different programming language but aiming to follow the same detailed logic (e. g., Meadows and Cliff 2012). One might reimplement the same model using a different modelling framework (e. g., Bagni *et al*. 2002). Or, finally, one might aim to mirror the model outputs, but using a significantly different model (e. g., Miodownik *et al*. 2010). These options represent a spectrum from “fidelity” (the same code on different hardware) to ability to demonstrate robustness in reported results (the policy implications are the same even if the models are significantly different).

In this article, we demonstrate the possibility and value of the last option, mimicking model outputs using a different model altogether. For clarity, to differentiate this option from the others, we describe this process as ‘reproduction’. In keeping with the practical importance of the approach, the article focuses on demonstrating its viability using a case study of the London School of Hygiene and Tropical Medicine (‘LSHTM’) model (Davies *et al*. 2020) which, along with the model from Imperial College (Flaxman *et al*. 2020), has played a key role in shaping UK COVID-19 policy advice.

In model reproduction, the aim is to design a model, the so-called Reproducing Model (‘RM’), based on the published descriptions of another model (the Target Model, ‘TM’) that mirrors the TM’s reported policy relevant outputs. In this case, we use an agent-based model (hereafter ‘ABM’) as the RM, but the approach is general and could also be applied to an ABM as a TM and, for example, a System Dynamics model as the RM. Reproduction can be coarse (e. g. the RM and TM both say that this intervention makes the death rate decrease) or fine (e. g. the RM and TM both say that this intervention makes the death rate decrease by 17.4%), but the aim in this article is to attempt reproduction and show that the approach is viable, what it involves and what benefits it can deliver.

The proposed reproduction strategy has a number of benefits:

- It is not dependent on potentially inaccessible detail and deliberately focuses on the published output from the TM, thereby maximising opportunities for community endorsement of model trustworthiness.
- It alerts us to the possibility that the results of a model may be dependent on its modelling technique (System Dynamics, Discrete Event Simulation, Agent-Based Modelling, network based, spatial or compartmental) rather than merely its assumptions.
- It draws our attention to the assumptions which cannot be inferred from the published research or may be contradictory across publications (and thus supports more transparency in model reporting).
- It serves as a potential robustness check on the extent to which the policy outcomes reported are the result of capturing general broad patterns of human behaviour rather than specific details of implementation and assumptions (or even programming errors).

As reproduction is an ambitious goal (the RM successfully reproduces key features of policy outcomes in the TM using published information alone) it carries a significant risk of failure: we might discover that we simply cannot reproduce policy advice from the TM. However, such a failure is not a negative result in scientific terms (and is actually valuable) because it implies that the TM as reported is inscrutable (given the skills of the researchers attempting to reproduce it) and may therefore be a poor basis for reliable policy. This is not to say that the results of the TM are wrong but that they cannot (in practice) be checked by anyone using public information.

The proposed strategy has advantages in its favour. Firstly, because models are complex objects, we are not obliged to be convinced (or sceptical) on the basis of reproducing a single policy intervention (which might happen more or less by chance). We can challenge the RM to perform on as many different policy outcomes as the TM generates and the better it performs overall, the more convincing we should find it and the TM. Secondly, because ABMs are particularly useful for experimentation and process analysis, trying to reproduce the TM may help us to learn more about what is being modelled. Thus, while the intended outcome of this approach is descriptive (matches well, matches weakly, or does not match), the process of achieving this outcome (even if negative) can still provide considerable scientific insight of wider value. We illustrate this claim in various arguments that follow.

The structure of the article is as follows. Section 2 describes core features of the LSHTM model (the TM) drawing on the published article and supplementary material of Davies *et al*. (2020). The third section describes mechanisms shared by the TM and our RM. The fourth section examines and accounts for the differences between the two models. The fifth section compares output from the two models to confirm the feasibility of the reproduction process. The sixth section justifies the claim that the process of reproduction is valuable independently of a successful outcome by delivering important new insights. The final section concludes by considering the limitations and challenges of reproduction.

## 2. The LSHTM Target Model

We describe the target model strictly as presented in Davies *et al*. (2020) to ensure that the account given here can be fully checked and criticised.^2^

Compartmental models (SIR, SEIR etc), of which the TM is an example, use ‘compartments’ which hold stocks of individuals in each disease state (for example S – Susceptible, E – Exposed, I – Infectious and R – Removed) and transitions between these states (Kermack and McKendrick 1927, May and Anderson 1991). The disease is modelled at the individual level as proceeding stochastically through a sequence of stages. At the outset all individuals (apart from those initially assigned to be seed infections) are Susceptible. Then, through Exposure (the detailed mechanism of which will be discussed shortly), they become Infectious. Infection may result either in having the disease asymptomatically (which is less likely to be transmitted and is more likely in the young) or having the disease symptomatically (after a period when one can transmit it but has no symptoms). The former infectious people are labelled Infectious-Subclinical (hereafter I_S_); they may infect others but display no symptoms. The latter become Infectious-Preclinical (hereafter I_P_), also infectious and asymptomatic, but then transition (“mature”) after some time into Infectious-Clinical (hereafter I_C_), where they display noticeable symptoms. Both of these routes lead ultimately to the Removed state in which the individual is either recovered, dead or in isolation (and, in each case, is assumed not to be able to transmit the infection further). The asymptomatic route lasts for a shorter time, is assumed to involve being less infectious, and is less severe, for example, never requiring hospitalisation. Figure 1 illustrates the overall structure of the disease state compartments in the TM and the transitions between them.

**Figure 1.**
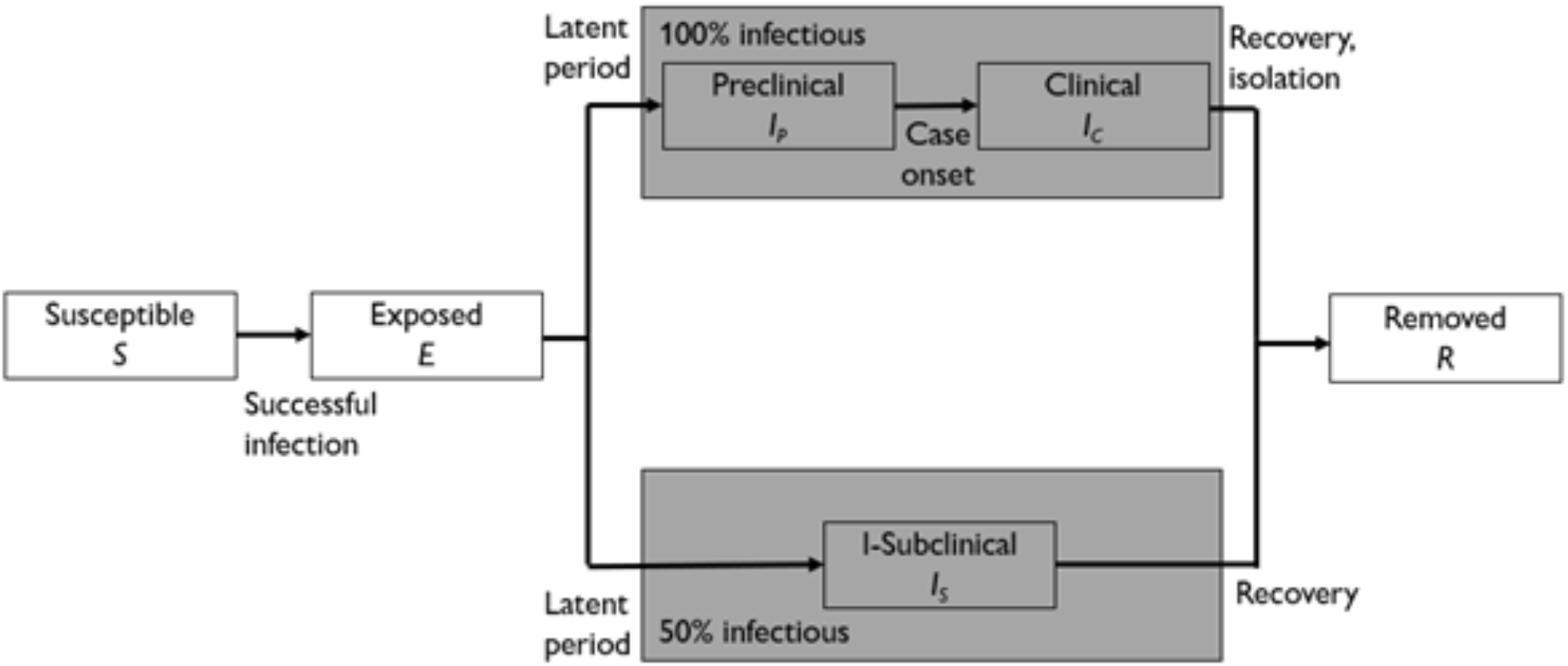
**The disease model in the LSHTM model showing disease states (boxes) and possible transitions (arrows) after Davies *et al*. (2020, p. e376)**

As well as being divided by their disease state, the simulated population are divided by age groups and administrative units (in this case UK counties). These subdivisions play different roles in the simulation, however. Members of one age group are in contact with, and can be infected by, members of any other age group. The rates of contact, as well as other rates such as the rate of showing symptoms, are age-based parameters supplied to the model. The age groups are used to capture empirically derived age-stratified contact matrices (a representation of the contact that each group has with every other), taken from the POLYMOD study (Mossong *et al*. 2008). For example, if a 50 year old worker has the disease and can transmit it (for example by going to work without PPE), then they have a high probability of infecting others of working age (e. g. aged 18 to 64) and a low chance of infecting other-household 5 year olds (who will be at home or school during the corresponding time periods). Data on contacts at home, at work, at school and in a composite “other” category (which includes activities such as travel and shopping) are used in the TM.^3^ The contact matrix data do not take account of the time of day or differences in contact rates at weekends.^4^

By contrast with age groups, counties are simulated as independent of each other in terms of disease transmission. There are no cross-border social contacts or infections, and hence each area must receive an initial number of seed infections in order to have an epidemic. Because this is the way that data is made available institutionally, Davies *et al*. (2020) use the 186 county-level administrative units in England, Wales, Scotland and Northern Ireland for their analysis. Each unit has data about its population stratified into the same 16 5-year age groups as the POLYMOD contact matrices (i. e., the number of individuals in each county from birth to 4 years, 5 years to 9 years and so on with a single group for those aged 75+). For each of the 186 counties an epidemic is simulated. Starting with the seeding of initial cases in each unit in the specified week (the first week for London and within the first three weeks elsewhere), the disease spreads between and within age groups (whether through contact at home, at work, at school or elsewhere) within the given county. The sum of individuals in different illness states across all the units gives the simulated total for the UK (something which could, in principle, be compared with the actual evolution of the epidemic).

To inform policy decisions, Davies *et al*. (2020) report UK total cases, peak cases, peak occupancy of intensive care unit (ICU) and non-ICU hospital beds, total deaths, and the time in weeks to the week with peak cases for different “policies” restricting contact. Clinical “cases” are defined in the model as new additions to the I_C_ compartment. To estimate the health burden (i. e., hospital beds occupancy) and the number of deaths from the disease, in each time step Davies *et al*. add proportions of the new cases to two parallel models. In the health burden parallel model, patients progress in and out of either the ICU or the Non-ICU compartment. In the deaths parallel model, they progress into a Deaths compartment. Neither of these parallel models influences events in the original transmission model (i. e. the compartmental structure of Figure 1). The parallel models are purely for generating policy relevant outputs.

The model is stochastic in its processes; it uses random sampling to determine flows between compartments and time spent in compartments. It also samples some of its parameters from distributions. Because of this random sampling, the results reported in Davies *et al*. (2020) are averages of 200 simulation runs. Each run is based on an *R*_*0*_ value sampled from a Normal distribution with mean 2.675739 and standard deviation 0.5719293. (These extraordinarily precise values are those quoted in Davies *et al*.) The need for sampling from a distribution reflects uncertainty about the actual value contextualised by empirical studies. *R*_*0*_ is the ‘basic reproduction number’ and is defined as the expected number of cases infected by one index case in a population where all individuals are susceptible to infection.

This framework creates a basis for the policy analysis that forms the bulk of the article. The analysis in Davies *et al*. (2020) covers multiple experiments, and, for brevity, we will not discuss them all in this paper. However, the first experiment is based around 6 policies, 5 of which modify contact rates in some way.

**Table 1.** The interventions described by Davis *et al*. (2020) relevant to the analysis in the present article.

| Policy | Effects on Contact |
| --- | --- |
| Baseline (No Intervention) | All contacts and infection levels are at 100%. |
| School Closure | School contacts are at 0%.<sup>5</sup> |
| Physical Distancing | Work and “Other” contacts (e.g. shopping) are at 50% of baseline. |
| Self-Isolation | The infectiousness of $I_c$ people is at 65% of baseline because those with symptoms stay home. |
| Shielding of Older People | Work and “Other” contacts are at 25% of baseline but only for those at or more than 70 years old. |
| Combination | School Closure, Physical Distancing, Self-Isolation and Shielding of Older People all in place simultaneously, each with the same effects on contact and infectiousness percentages as before (but merged). |

After running the baseline scenario, the TM identifies the date at which the peak in UK cases occurred during that run. For each intervention scenario (2-6) the TM schedules the relevant changes to the contact rates and infectiousness to begin 6 weeks prior to the peak cases found for the baseline.^6^ The end to the intervention is scheduled for 12 weeks after it starts. At this point, parameters are returned to 100% of their baseline values.

This summary of the LSHTM model will now form the basis for describing the RM in the next section. The similarities and differences between the models and analysis of the success of the reproduction process (both directly and in terms of what was learned from carrying it out) then follow in subsequent sections.

## 3. Mechanisms Common to the TM and RM

The reproduction model (RM) was coded in NetLogo (Wilensky 1999) as an agent-based model, with the agents representing individuals and their associated interactions with each other in regard to disease transmission within an “environment” (for example being in the same physical location at the same time).^7^

Both TM and RM simulate people passing between compartments representing disease states. For the purposes of evaluating interventions, the TM defines everyone passing from I_P_ to I_C_ as a new recorded “case”. For reasons discussed in the next section, our model (shown in Figure 2 below) differs slightly in compartmental structure from the TM. During the transition from I_C_, our cases divide, with some going straight to a state called “Recovered”, a second proportion going to Intensive Care Unit (ICU) beds in hospitals and a third group to non-ICU hospital beds. After the two hospital bed states, patients transition either into the Recovered state or into the Death state. All I_S_ people transition into Recovered. Recovered people are immune and not infectious, but the possibility of immunity wearing off over time and their transition back into Susceptible would be a simple addition to a later version of the model. Death is a terminating state (which means that agents can never leave it subsequently). Following the TM, we assume I_S_ individuals are only half as infectious as those in the other infectious states.

**Figure 2.**
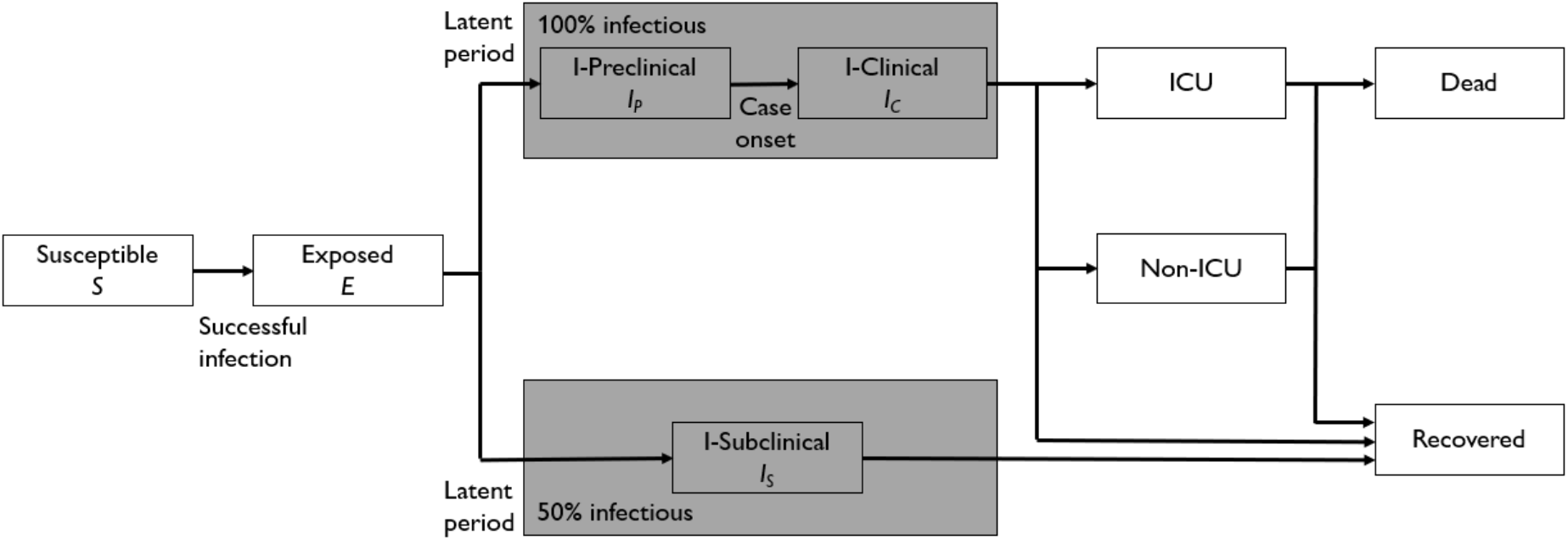
The disease model in the Reproducing Model (RM) showing disease states (boxes) and possible transitions (arrows)

The simulation is initialised by the creation of an agent population. While the TM simulates each of the 186 UK county-level administrative units and then sums the resulting 186 time series to make an aggregate time series for the whole country, our ABM simulates only a single county, Rutland (the smallest county that is not an island) with 39,697 people. The reason for this is that, given the assumption of the LSHTM model that there is no cross-boundary infection (and that the separate individual county epidemics are therefore simply added up) nothing is added to our reproduction attempt by modelling the whole UK (and it is obviously far more computationally demanding.) Our population of agents is generated with the same ages and genders as the people in the demographic data file used in the TM for Rutland. On first initialisation, all agents are formally assigned to the Susceptible state (though some are then transferred to the Exposed state almost immediately as seed cases).

Data for contact matrices, parameters defining the interventions, the dates of UK school holidays and the *R*_*0*_ values are all taken from the TM. As in the TM, interventions are defined by the percentage reductions they are assumed to create in contact rates, and by the reduction in infectiousness of I_C_ self-isolating individuals (scenarios 4 and 6).

After initialising the clock time to 0, representing 29 January 2020 (the date of the first confirmed UK case), various time-based events are scheduled in both TM and RM. Potentially infectious social contact events are scheduled to occur every 6 hours. Also processed at 6 hour time steps in the TM are transitions between disease states. (The RM is more flexible in its timing of state transitions, as detailed in the next section.) An intervention scenario is scheduled, in which the contact rates and relative infectiousness of I_C_ are modified (as noted in section 2) at the start of the intervention, and are returned to the baseline values at its end (12 weeks later as already explained). Seed infections occur using the same schedule as in the TM for Rutland: two people per day for 28 days, beginning on the same day as used in the TM, which was a randomly chosen day in each run, within the first three weeks of the simulated period. Finally, the calculation of output statistics and the consequent updating of charts are scheduled for midnight on each simulated day.

For contact events, following the TM, we process all Susceptible agents in sync. For a Susceptible agent in age group *i*, a measure of their overall exposure risk (“force of infection”) is calculated using Equation 1.

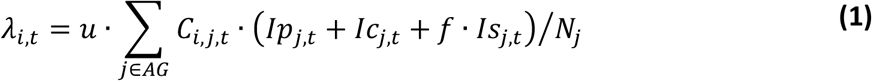

AG is the set of 16 age groups. *C*_i,j,t_ is the number of contacts per day at time *t* between someone in age group *i* and people in age group *j. Ip*_j,t_, *Ic*_j,t_ and *Is*_j,t_ are the numbers of people in age group *j* in each infectious state^8^ and N_*j*_ is the total number of people in age group *j. f* is the relative infectiousness of asymptomatic I_S_ people, set to 50%. The susceptibility parameter *u* represents the probability that a contact with a fully infectious person results in infection. *u* is related to *R*_*0*_ by the next generation matrix, whose *i*^th^ and *j*^th^ cells are given by:

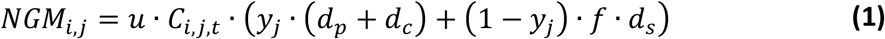

where *d*_*x*_ is expected transition time from infectious state *x* (p, s or c), and *y*_*j*_ is the symptomatic rate for someone in age group *j. R*_*0*_ is the absolute value of the dominant eigenvalue of the next generation matrix. This completes the calculation of the force of infection, the rate at which potential infection events occur to a susceptible person. Given the rate λ _i,t_, the probability that a susceptible person will be infected during a six-hour time step is:

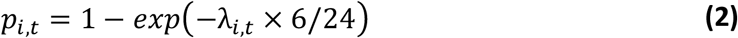

Given this probability and the number of susceptible people in age group *i*, the number of people being infected in the current time step is randomly sampled from a Binomial distribution. The ABM then has to randomly sample which of the susceptible agents they are and processes their transition to the Exposed state.

Upon determining their next disease state other than Susceptible, a person has their transition (‘maturation’) event out of that state scheduled. Times to maturation (also sometimes called maturity) for each state are sampled from Gamma distributions, with parameters taken from the TM. Where multiple routes lead from a state, the next state is determined using parameters based on age group (again provided from the TM). Examples of these branch rates include the symptomatic rate, hospitalisation rate, intensive care rate and death rate.

## 4. Differences between TM and RM

Our agent-based model implementation (the Reproduction Model, RM) differs from the LSHTM Target Model (TM) in several ways. Many of these reflect the different styles of modelling involved (compartmental versus ABM) and therefore if we can still reproduce the policy outcomes of the TM using the RM, this would support the contention that these might be substantive behavioural outcomes and not mere artefacts of the modelling approach. In the first two subsections we describe two *potential* differences between the models. The ABM offers greater flexibility in representing agent heterogeneity and timing of events, but for the purposes of reproduction, we restrict ourselves to the heterogeneity and time schedules of the TM. The remaining subsections, by contrast, describe changes forced upon us by the need to explicitly represent each agent undergoing a coherent sequence of processes in an ABM.

### Representing agent heterogeneity

The TM represents all people in the same disease state compartment, transition time sub-compartment (representing how long before they mature or move to the next state) and age group by a simple count. In this modelling approach, people have no other attributes. Further characteristics (e. g. key worker/non key worker; compliant/non-compliant with mask wearing) would require a further sub-division of compartments and additional parameters. In the RM, the attributes of each individual agent are explicitly represented. To replicate the TM, we only need to use age (for parameters based on age group), current disease state and the time of the next state transition. But a person’s gender, family relations, household cohabitants, school peers and co-workers could all straightforwardly become relevant in an extended model (as they almost certainly are in reality and in network-based approaches to disease transmission – Eubank *et al*. 2010). For now, contact rates in the RM are based on age groups alone and contact partners may be anyone in the same category (for example “workers”). In order to reproduce the TM, the RM currently does not represent repeated contact with the same partners, such as spending every night with your family or most days with the same co-workers, although an ABM readily allows this functionality and it seems likely to be empirically important.

### Time

The TM does all updating, both infections and other state transitions every 6 hours. Transition to the next compartment is governed by a time priority queue for each disease state and age group sub-compartment. This means that the model already “knows” when someone is going to make the next transition before they actually do so (but no behavioural implications follow from this). The RM implements a three-phase discrete-event simulation (DES) engine (Tocher 1963), giving it the potential to model state transitions occurring “between” the current 6 hour time slice updates (and each agent does not have to transition simultaneously with others). The current RM maintains the TM’s schedule for computing infections, so there are no behavioural consequences from the difference between RM and TM in state transition synchronicity. But the DES engine allows for future extensions of the ABM. For example, if contacts occurred within particular locations (workplaces, schools or homes for example), agents could change locations according to their own personal schedules, rather than in sync at common time steps. Furthermore, the DES engine allows the introduction of more precision in contact and transition times, without significant increase in computational work. To obtain more precision in timing from the TM, by contrast, would require simulating more time steps per day which is obviously more computationally intensive.

### Verification and validation at the micro level

In the RM we can trace an individual agent through its disease states. This is not possible in the TM, where we can observe only aggregate numbers of individuals in sub-compartments. Each such number is the result of some people flowing into the sub compartment, others flowing out, and some remaining (while not yet “due” to transition through maturation). Given only the numbers in each compartment at a specific time point, it is impossible to establish how many agents have transitioned and where. For example, if compartments A, B and C each initially have 10 people in them (and in the next two time steps have 9, 11 and 10 and 9, 10 and 11 respectively) we cannot tell if one person has just jumped from A to B to C in two time steps or whether someone has gone from A to B, followed by a different person going from B to C. There is thus no personal history to record, despite the TM being described as an “individual-based model”. In an ABM, agents can record their state transitions, allowing us to analyse (and thus potentially validate) the distribution of total time spent infectious or ill (which can serve as another potential source of validation against individual “biographical” data distinct from aggregate disease states).

### Compartmental structure

Once one considers the personal history of an agent, it becomes apparent that an ABM will have to restructure the compartments of the LSHTM model. The TM has a SEI3R^9^ structure to simulate the transmission of the disease, where R stands for Removed from being infectious, rather than Recovered (and therefore also includes those in isolation which is assumed to be totally effective). It adds parallel processes (based on “cloned” populations) to this transmission model to simulate the health burden (occupation of hospital beds in ICU and non-ICU categories) and deaths resulting from I_C_ cases. The flow of people from I_P_ to I_C_ is thus cloned twice. One clone flow is fed into ICU and Non-ICU compartments to assess health burden. The other is used to establish the death toll. Depending on the randomly sampled maturation times for transitions between compartments, it is possible (though rare), for the TM to be adding one to the death toll for a person who is also still registered in a health burden compartment and in the I_C_ compartment. Within the context of the TM, this technical oddity has no problematic behavioural implications because it can have no bearing on infection rates (at least based on other model assumptions about disease transmission in hospitals) and the interest of Davies *et al*. (2020) is in UK averages rather than micro-level variations.

For an agent-based approach this order of events is not coherent. Disease states must occur strictly in the sequence: become ill, go to hospital, and die. For this reason, we were obliged to merge hospital and death states into the transmission structure and adjust mean maturation times to best match the corresponding timings in the TM (as the fairest achievable basis for comparison between the two models). Figure 2 (which can be compared with Figure 1 for the TM above) shows the overall structure of disease compartments and transitions for the ABM.

The Removed compartment has been replaced. The health burden (ICU, Non-ICU hospital beds) and death parallel processes have been merged into the main structure, with a new compartment added for Recovered. For reproduction purposes, we still assume that people are removed from being able to transmit once in hospital (or the morgue), though future extensions could add hospital compartments to the force of infection equation, down weighted to reflect the (hopefully) much reduced risk of transmission there.

Restructuring the compartmental model brought to light issues concerning the parameters for transition and branching between states. Firstly, rates have been derived from studies using age bands that differ from the 16 in the POLYMOD contact matrices (and so have to be converted in the TM code). This conversion is not documented in Davies *et al*. (2020). Secondly, to generate deaths the TM code uses a Case Fatality Rate (CFR), which is confusingly labelled in the code “prop_noncritical_deaths”, even though it applies to all “cases” and not just “noncritical” (i. e. Non-ICU) patients. For our restructured RM we need fatality rates for people in ICU and Non-ICU states, rather than for all clinical cases. Dividing the CFR by the hospitalised rate gives the same number for every age group, 12.4%. Even though our modelling approach would easily allow us to give ICU and non-ICU patients the distinct chances of dying likely in reality, lacking further data we decided to use the same hospital death rate, 12.4%, for both ICU and Non-ICU patients. Although this is unlikely to be true empirically and may, in any event, be assumed mistakenly in Davies et el., we nonetheless follow their specification.

Another difference required by the distinct modelling approaches of the RM and TM was in our choice of transition time distributions. In the TM’s parallel processes, patients flow straight from I_P_ to Death, as well as to ICU and Non-ICU (see Figure 1). In the RM, the routes to ICU and Non-ICU are via I_C_, and those to Death (and also Recovered) are via ICU or Non-ICU (see Figure 2). So, in order to follow the logic of model reproduction, the transition time distributions associated with each state transition must have different parameter values in the two models. Ensuring that the mean total maturation times are correct is simple arithmetic. Since the different transition processes are independent of each other, they can be summed.

One final change concerned when the TM and RM sample transition times. For TM, they are sampled for each individual entering a compartment. If there are multiple routes out (as there are for Exposed, and for I_P_), the number going down each route is sampled on *exiting* the compartment. Hence the TM assumes that people progressing to I_P_ and I_C_ spend the same length of time between becoming infected and becoming infectious, as those people progressing down the asymptomatic subclinical route. This may or may not turn out to be true of COVID-19. For the RM, an agent’s route out is sampled on entering the compartment (i. e. at the time of infection, the RM decides whether the agent will ever become symptomatic) and a time to reaching their next compartment is then sampled. Thus, transition times to different states can follow different distributions. This does not matter for the routes out of Exposed – at present, no one knows whether incubation times differ significantly between future symptomatic and asymptomatic individuals, and it would be hard to gather empirical data to address this, given that we rarely know exactly when infection occurred. But for the routes out of IC it could be important. Time in IC is time spent still making social contacts with Susceptibles. Those clinical cases requiring hospitalisation (transitioning to ICU or Non-ICU) may differ in this amount of contact time from those clinical cases passing straight to Recovered without the need for a spell in hospital.

### Stochasticity

TM and RM are both stochastic models but require different numbers of random samples. In the TM, stochastic flows from Susceptible to Exposed are simulated by sampling from a Binomial distribution, with parameters *n* (the number of Susceptible people in the age group) and *p* (the transmission probability derived from equation 1). Similarly, branching (such as from Exposed to either I_P_ or I_S_) is decided by sampling from Binomial distributions. The RM also uses Binomial distributions to determine *how many* people in the current time step are becoming infected or flowing down a particular route. But as an ABM, it then needs to determine *which* people they are. For example, if a Susceptible compartment contains 100 people, both TM and RM sample one Binomially distributed random number to determine how many (say 10) are being infected at that time step. But the RM then needs to compute the first 10 items of a random sequence (which requires sampling 10 uniform random numbers) to determine which of the 100 are becoming Exposed.^10^

Only in the case of maturation times do TM and RM sample similar quantities of random numbers (as in this respect TM acts as an *individual-based* simulation). Gamma distributed times are sampled for each individual in both models. A simulation in which most of the UK population became infected would need tens of millions of Gamma distributed random numbers per simulation run for each compartment transitioned from.

### Population

An ABM that simulates the entire UK population, whether directly or by simulating counties, will make heavy demands on computer time and memory.^11^ Partly for this reason, we decided to restrict the reproduction attempt to a single county (Rutland, the smallest county that is not an island). In the RM we therefore filtered out all other counties from the TM’s data input files.

For some outputs, choosing a small county should not affect the comparison. In common with other compartmental models, stock levels (e. g. total and peak cases) scale well with population, except for very small populations (for example 100 people), where stochastic variation can have a significant effect on outcomes.

However, time to occurrence of various events (such as the time to peak cases and time to ICU beds reaching full capacity) prove to scale instead with the logarithm of population divided by the number of seed infections (Watts *et al*. 2020). Hence, extrapolating times from studying a small county to the UK population (66.4 million) is less straightforward than extrapolating compartment stock levels and we therefore did not attempt this. As Watts *et al*. (2020) note, supplying the same number of seed infections to each county means that their times to peak cases vary with the logarithm of their sizes, which raises problems when aggregating their time series to simulate the UK (as the TM does). The county peaks will tend not to be in sync, and consequently when these are summed to produce the UK aggregate, its peak will be lower than if the counties had been in sync. For the purposes of comparing the models most effectively through reproduction, it is thus advisable to focus on one county. Whether Davies *et al*. (2020) were right to assume they could simulate the UK as an aggregate of independent counties is an issue for further debate.

### Parameters and dates

For our experiments (see the next section for results) we chose to *import* into the RM some parameters and dates *generated* by the TM, rather than reproducing the TM’s generative mechanisms.

Where the TM generates an *R*_*0*_ value for each simulation repetition by sampling from a Normal distribution, we modified the TM code to export these values. We then imported 50 of them into the RM, so that we could compare model outputs for the same *R*_*0*_ values. We chose 50 *R*_*0*_ values evenly spaced across the possible range (rather than the full 200 used in Davies *et al*. 2020), for reasons of computer time, clarity of graphical presentation and owing to the fact that in practice the 50 cases seemed sufficient to demonstrate the good match between TM and RM.

Also exported from the TM were dates of seed infections and dates of beginning interventions. Again, by reusing these dates in the RM, we ensure a close comparison.

Both models used the same age-based demographic data to generate the population for the county of Rutland, the same rates of hospitalisation and death, the same list of UK school holiday dates, and the same contact matrices. These are either imported to the relevant model or hard-coded. However, the age-based symptomatic rates are sampled in each simulation run from a file generated from an earlier attempt by Davies *et al*. (2020) to fit a transmission model to some COVID-19 data using Markov Chain Monte Carlo techniques. The file contains a Markov Chain estimating the joint posterior probability distribution for sets of age-based symptomatic rates. Sampling from this distribution reflects the degree of uncertainty about symptomatic rates (and thus the variance in – or uncertainty about – model outputs given this uncertainty in input). Rather than export the actual symptomatic rates used in each run of the TM, the RM samples from the same Markov Chain. In this way, what we reproduce is the TM’s degree of uncertainty about symptomatic rates (rather than the particular rates sampled).

To reiterate our argument, the aim was to make the RM as close to the description of the TM as compatible with our different modelling approach (agent-based rather than compartmental) and, wherever possible, to make assumptions that were merely technical with no behavioural effects that would create anomalies (like people dying and then recovering) or significantly disrupt the logic of the TM. “Significant” is defined here by the ability to reproduce the TM using the RM. Small disruptions of logic that still allow reproduction may actually vindicate the *robustness* of the TM results, as argued above. The test of our claim to reproduce (rather than replicate) the TM remains our ability to track its policy outcomes (rather than intuitions about the plausibility of our approximations). It is to this test that we now turn in the following section.

## 5. Comparing the TM and RM

Where it has not been purely descriptive, the argument to this point has been entirely conjectural. *If* we can reproduce the TM from published research using the RM, then this outcome (and the process that achieved it) will be useful. The aim of this section is therefore to show that we *can* in fact do this. Figures 3 to 5 demonstrate the extent of reproduction for three measures of the TM (total cases, total deaths and time to the peak number of cases) in both the baseline case and under the “combination” intervention described above. The blue dots in the Figures represent the values from the TM. For each *R*_*0*_ value, the RM was run for 100 repetitions. Crosses mark the median run from the RM, and the two ribbons indicate 50% (from the 25^th^ to the 75^th^ percentiles) and 95% (2.5^th^ to 97.5^th^) prediction intervals.

**Figure 3.**
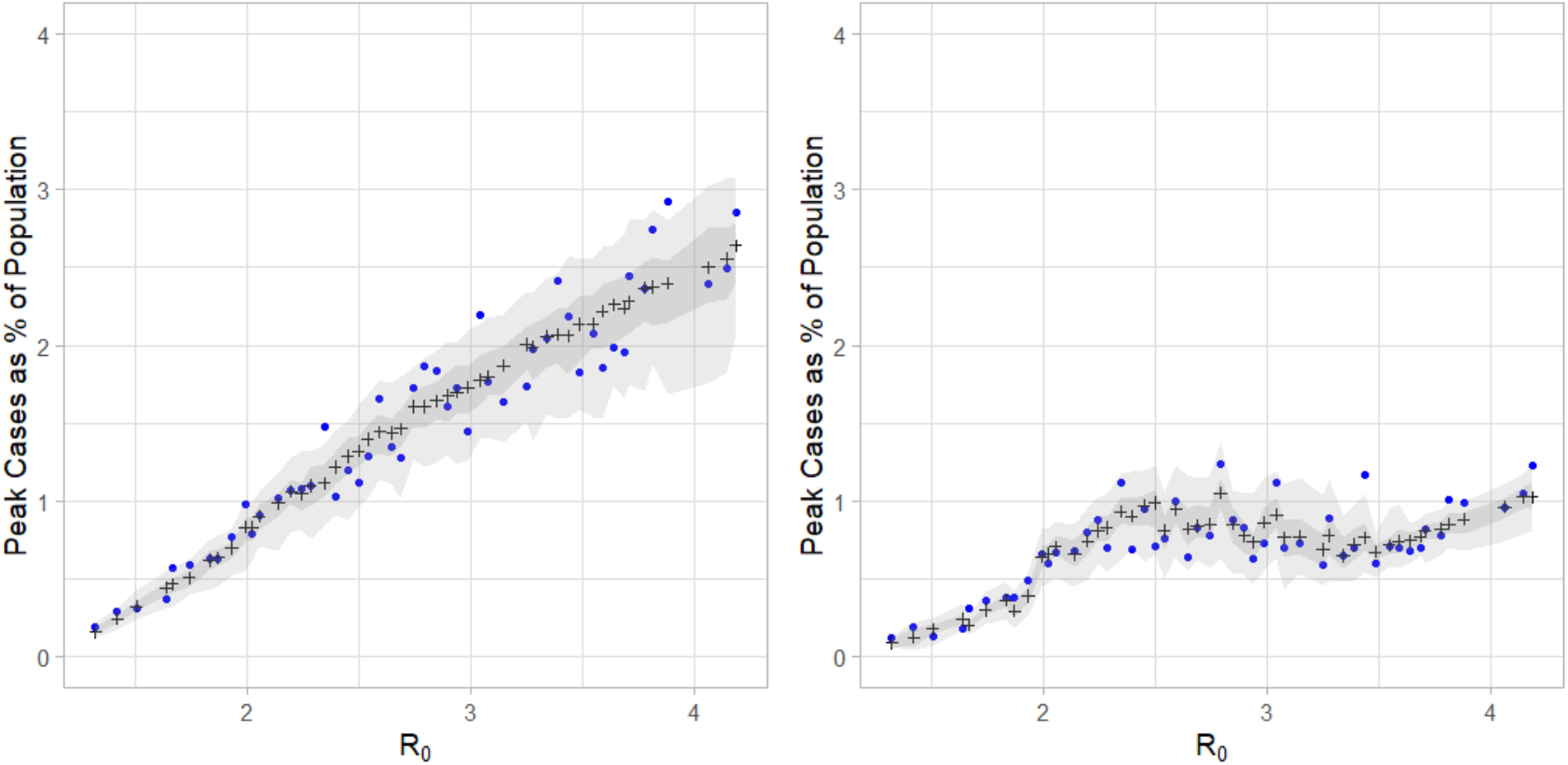
**Comparison between TM and RM outcomes on the measure of “peak cases as a percentage of population” for the baseline simulated epidemic (left) and the “combination” intervention (right).**

Figure 3 shows that the RM is able to mirror both qualitatively and quantitatively what happens to the peak in cases when the “combination” intervention is introduced relative to the baseline. However, the reproduction is somewhat imperfect since the target data from the TM is outside the 95% prediction intervals from the RM surprisingly often and also appears to be found mainly at the edges of the ribbons, rather than more clustered around the medians.^12^ Nonetheless, this data is quite persuasive as a justification for developing the reproduction approach further.

Figures 4 and 5 support the same conclusion (that the RM can do quite a good job of tracking the effect of policy interventions in the TM both qualitatively and quantitively).

**Figure 4.**
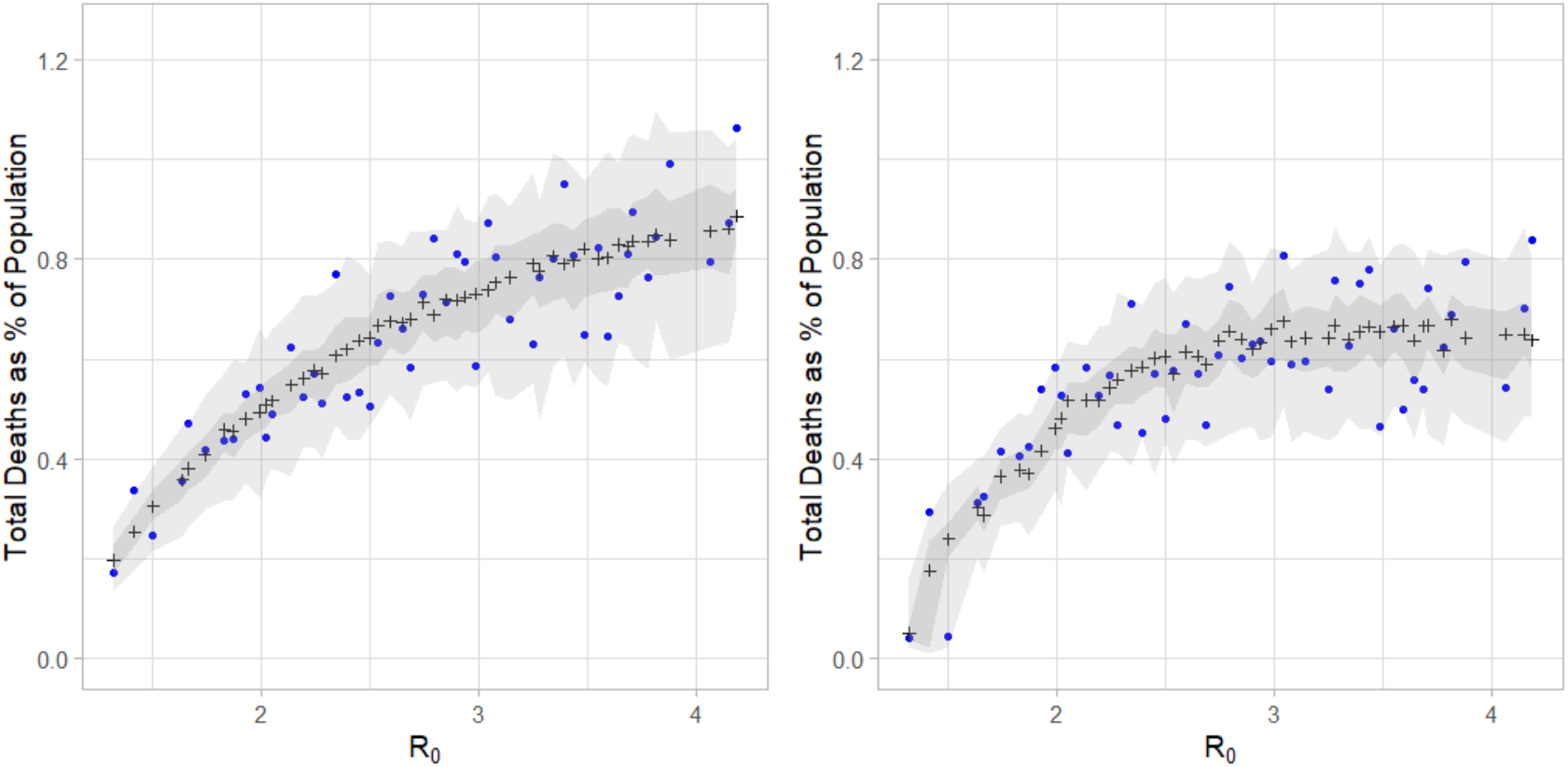
**Comparison between TM and RM outcomes on the measure of “total deaths as a percentage of population” for the baseline simulated epidemic (left) and the “combination” intervention (right)**

**Figure 5.**
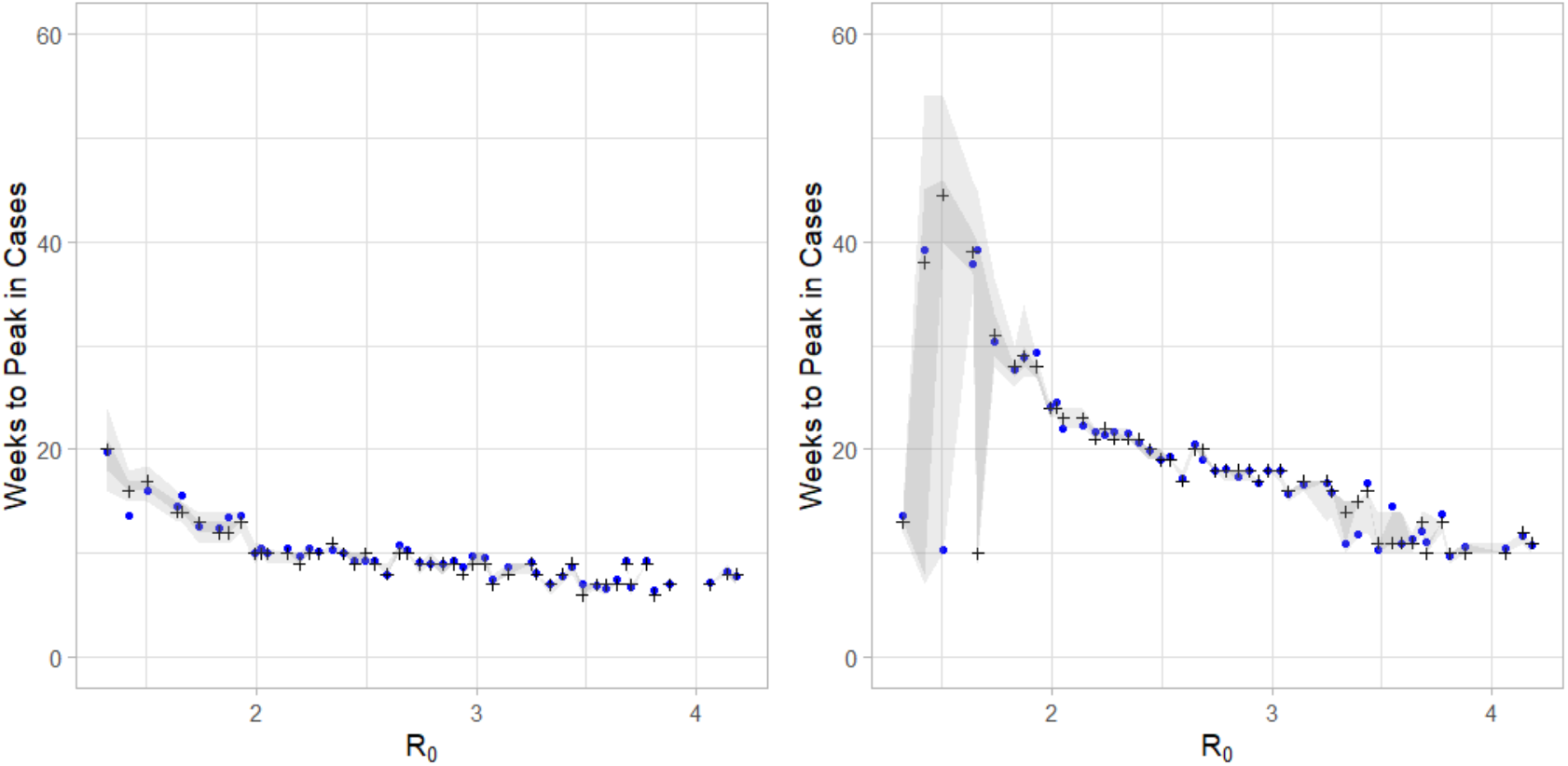
**Comparison between TM and RM outcomes on the measure of “weeks to peak in cases” for the baseline simulated epidemic (left) and the “combination” intervention (right)**

It is interesting that the RM has managed to mirror the apparent bifurcation of weeks to peak cases under the combination intervention when *R*_*0*_ is low. This phenomenon appears to *emerge* from the behavioural assumptions of the TM and RM (and is not specifically reported by Davies *et al*. 2020) so the ability of the TM also to display it as an emergent phenomenon provides good evidence for the claim that we have successfully reproduced the LSHTM model.

## 6. Lessons Learned from the Reproduction Process

The reproduction process provides benefits for both the TM and the RM. If the RM reproduces the TM, then it adds independent credibility to the TM’s policy prescriptions. If the RM does not reproduce the TM, it warns us that the TM may be erroneous (at least based on the information about it that has been made readily accessible.) However, in addition (as already suggested), regardless of whether reproduction is successful the process of attempting provides further understanding of the TM, and may be useful for the authors of the TM to update and improve their model and documentation.

### Co-verification

The aspiration of reproduction is that it should succeed from the academic paper alone. In fact, this did not prove possible for the LSHTM model: we had to have recourse to the publicly available codebase deposited by Davies *et al*. on GitHub. Nonetheless, the focused aim of reproduction made it much more feasible for us to learn effectively from the code and documentation because we knew what issues we were trying to address (rather than trying to make sense of all the code from scratch.) The idea of co-verification is to recognise that when TM and RM do not match, the problem could lie with either model. The process involves debugging in the usual way but going back and forth between the models (while comparing their behaviours) to establish which is more likely to be at fault.

This point is nicely illustrated by what we discovered because the match between TM and RM reported in the last section is not what we initially found. As Figure 6 shows, the initial attempt at reproduction was a dismal failure but the process of comparison also identified the nature of the problem and therefore suggested precisely how we could check and thus resolve it.

**Figure 6.**
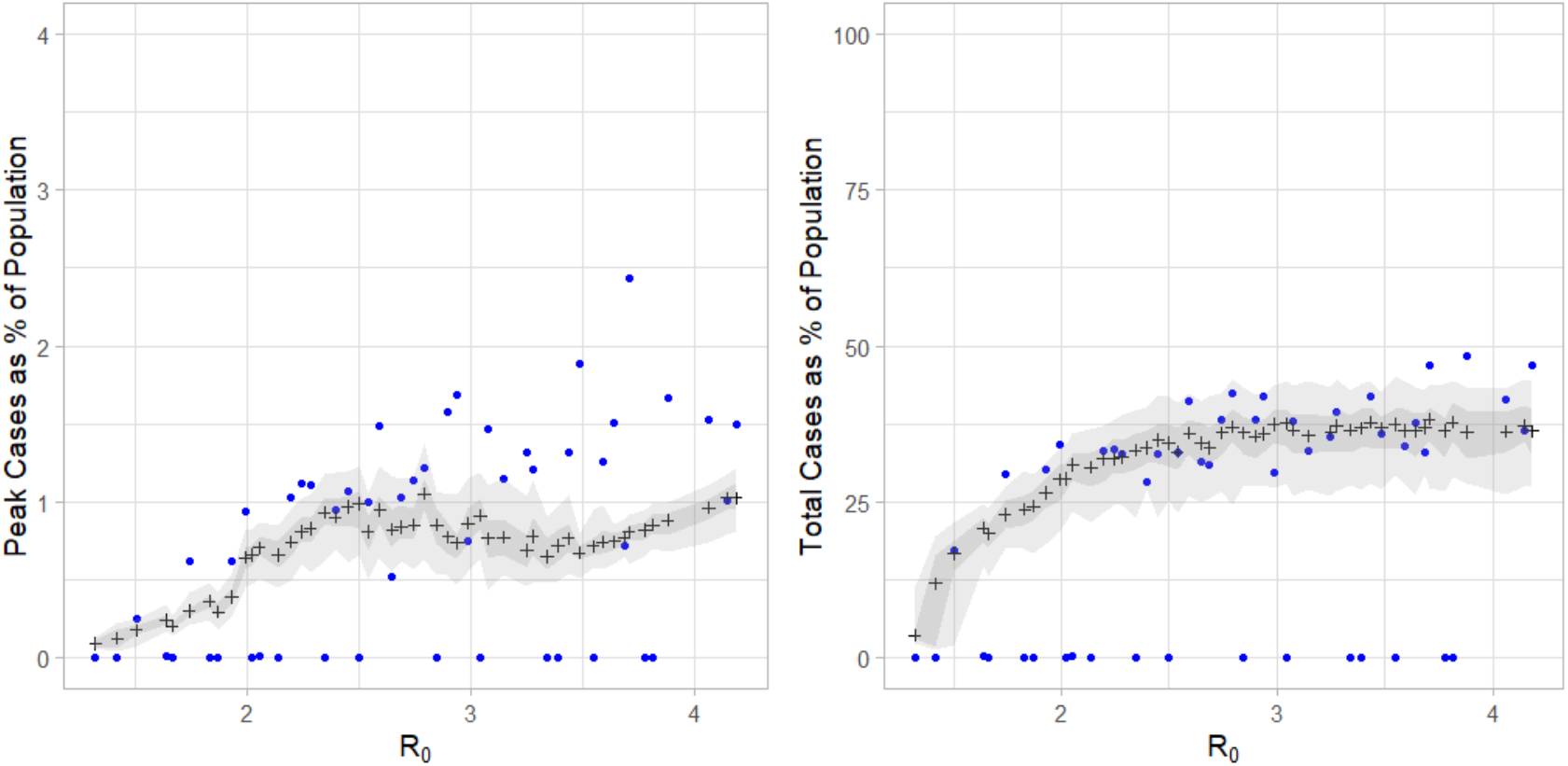
**Relationship between RM (grey crosses and ribbons) and TM (blue dots) for two model measures (“peak cases” – left and “total cases” – right) under the “Combination” policy and *before* the co-verification process described in the text. The crosses are the median results from the RM and the ribbons are the 50% (dark) and 95% (pale) prediction intervals.**

As Figure 6 clearly shows, some simulation runs of the TM were simply not resulting in epidemics. This unexpected result could therefore be traced with relative ease to a bug in the part of the TM code dealing with the seeding of infections.^13^ Fixing the bug involved ensuring that intervention scenarios used the same seed infection schedule as the baseline scenario (and with this correction alone we obtained the convincing reproduction reported in the previous section thus confirming the hypothesis that we had discovered the source of the reproduction failure).

*Comparison* of specific models thus gives the modeller a more effective “place to stand” in deciding whether it is the TM or RM that is more likely to be at fault (and thus where to look more precisely to correct the issue). In this case, the “failed epidemics” were made more conspicuous both by their absence in the RM output and by the reproduction aim of comparing the *distribution* of individual simulation runs (rather than the aggregates reported in Davies *et al*. which presumably had obscured the “failed epidemic” runs.)

### Robustness testing

One way of thinking about different modelling approaches is in terms of the assumptions that they take for granted or find it difficult to give up (for example on technical or analytical rather than empirical grounds.) Reproduction, which can be attempted using different modelling approaches (as here with an ABM reproducing a compartmental model) can give insight into the robustness of model results based on assumptions that are not questioned or possibly cannot be relaxed within a particular modelling approach. An example of this is provided by previous research involving the authors and collaborators (Watts *et al*. 2020). This drew attention to the sensitivity of compartmental models to both population sizes for the units of analysis (an epidemic in Kent behaves different from an epidemic in Rutland but the results of county epidemics are nonetheless aggregated in the policy analysis presented by Davies *et al*.) and to the exact seeding assumptions (which of necessity are rather weakly grounded in data, the whole issue with most diseases being that by the time you realise someone has got it, they have probably already passed it on.)

Clearly the argument in Watts *et al*. (2020) stands on its own intellectual merits but the point for the present article is that, as with the co-verification example above, this issue was bought to light by our attempt at reproduction. Specifically, it was the practical need to model Rutland (rather than the whole UK) that focused our attention on unit population sizes (and their consequences) and the oddity of “failed epidemics” which made us aware of the sensitivity of the TM to seeding assumptions.

Obviously, it is not appropriate for us to reason about what Davies *et al*. did *not* say but we can report that (for whatever reason) they did not present sensitivity analysis about seeding assumptions or discuss the implications of aggregating differing population unit sizes (both of which we have shown to matter non-trivially to model outcomes and thus *ex post* to merit some justification). The point is not just that we have found a potential vulnerability in the robustness of the results presented by Davies *et al*. but that, without the reproduction process, we would not have known where to focus our search for such vulnerabilities (with an *undirected* sensitivity analysis being potentially very resource intensive.)

### Surfacing assumptions

This brings us to the final class of learning we achieved in carrying out the reproduction reported here. Trying to reproduce the TM made several of its assumptions much clearer and thus raised possible concerns about the relations between their implications and reality. For example, as already suggested, the LSHTM model creates separate (and independently seeded) “county epidemics” and then aggregates them to produce a projection for the UK as a whole. The “failed epidemic” bug confirmed that when there was “inadequate” seeding, no epidemic would occur in a given county. The implication of the model is therefore that nobody ever crosses county boundaries! However, this issue is a little less obvious than it may initially appear. If each county epidemic is seeded at more or less the same time and with the same “strength” (as Davies *et al*. specify), then whether or not there is assumed to be cross country movement, its effect will be swamped in practice by exponential infection growth through the normal operation of the compartmental model. Thus, in a sense, one assumption becomes irrelevant (and thus less visible) in the face of another. If, on the other hand, the assumption of synchronous seeding of equivalent strength is not empirically supported, then the absence of cross county movement (which clearly occurs to a considerable extent in the real world^14^) ceases to be a legitimate simplification. This concern is compatible with freely available data on the earliest confirmed cases and the first recorded cases in some geographical units. For example, the earliest cases were not in London boroughs (which the LSHTM model seeds slightly earlier) but in York and then Brighton and the first cases were not confirmed in Wales, Scotland and Northern Ireland until the 27^th^ and 28^th^ February (already at the very edge of the 4 week seeding window assumed by the LSHTM model for units outside London.) This data, which is sparse but nonetheless reliable, is compatible with the reasonable idea that travel between counties and differential infection rates (for example through airports and their connections to particular towns) would also contribute to an explanation of the geographical distribution of COVID infections over time. Given the limited behavioural role of counties, it seems unlikely that epidemics are independent as the LSHTM model assumes.

It is important to be clear that we are not saying that our conjectured assumptions are “right” or even “better” than those presented in the LSHTM model (though we have at least suggested what data could be used to explore this matter namely dates of early cases by county) but without being surfaced and therefore having at least the potential to be evaluated in the light of data, any dialogue about model assumptions cannot even begin.

To sum up then, even if we had *not* succeeded in reproducing the TM, we would still have learned several different kinds of lessons valuable to evaluating the LSHTM model and its policy recommendations.

## 7. Conclusions

This article demonstrates, at least provisionally, the feasibility and value of the reproduction process proposed. It *is* possible to reproduce the outputs of one model by developing another model, if not entirely then rather largely, on a published account of it. This process not only has the potential to endorse independently the reliability of the TM for policy (if it works) but to raise concerns about its suitability as a policy model (if it does not).

The replication of the TM was successful (after we had corrected a seed infections bug in the TM) across a range of values for *R*_*0*_, displaying outputs for cases, hospital beds, deaths and times to peaks very close to TM in median values and deviations from typical behaviour. There are slight differences in the degree of dispersion around typical behaviour but any difference is too small to alter the policy recommendations of the model.

The approach thus deals with an important concern within the larger scientific community about the “inscrutability” of models used for policy within the larger scientific community (which is generally tasked with ensuring research quality but may find it particularly hard to do so in the field of modelling.) But even in the event that a match cannot be achieved, we have also shown how attempting reproduction can give important insights into the workings of the TM and the relevant social processes (in this case of non-pharmaceutical epidemic control). We think the quality of match (which was achieved while the pandemic was ongoing by a small team working informally) is easily sufficient to justify developing the reproduction approach further.

Our ability to replicate the TM in this case is a testament to its quality. Without sufficient documentation (whether in the academic article or supplementary code), the model reproduction process would be impossible. However, with a high quality TM, the benefits of the model reproduction process are clear.

Given the potential value of the approach, we were keen to make the work available as soon as possible. This being so, it is clear that the reproduction process could have been taken further (comparing more policies, attempting to improve the quality of match so that the RM data were more convincingly within the TM prediction intervals and so on.) However, we also had a broader agenda in developing the reproduction approach which was to examine the general robustness of compartmental models to their assumptions (particularly those which are less visible). In practice, we had to considerably restrict the representational potential of the ABM to achieve reproduction. Now that we have an ABM capable of tracking the LSHTM model convincingly a more interesting question for us is whether its results would still hold if it did not make the sort of assumptions typically made by compartmental models. It seems to us that different approaches to modelling, all of which capture certain aspects of reality but tend to downplay others may be problematic when compared to the possibility of a coherent synthesis.

The final point is what we can (and perhaps should) learn from the experience of COVID. The most effective response from the modelling community to the next pandemic (which is almost inevitable in statistical terms at least) is not a proliferation of unassessed models once the disease has already broken out but an ongoing devotion of resources to an infrastructure which supports improving model quality and is congruent with the kind of “competitive cooperation” found in good science. It is to this aim, whose value should value obvious, that this paper hopes to contribute. We cannot oblige politicians to attend to our judgements of which models can be trusted but that does not mean that we should not attempt to establish it.

## Data Availability

https://github.com/innovative-simulator/CovidModelAgentBasedReproduction

## Appendix

**Table A1.** Intervention scenarios and their modifications to each contact matrix and the relative infectiousness of people in the I_C_ state.

| Scenario | Home contacts | Work contacts | School contacts | Other contacts | Infectiousness of $I_c$ individuals |
| --- | --- | --- | --- | --- | --- |
| 1. Baseline | 100% | 100% | 100% | 100% | 100% |
| 2. School Closure | 100% | 100% | 0% | 100% | 100% |
| 3. Physical Distancing | 100% | 50% | 100% | 50% | 100% |
| 4. Self-Isolation | 100% | 100% | 100% | 100% | 65% |
| 5. Shielding of Older People | 100% | 25% (age $\geq 70$);<br>100% (age $< 70$) | 100% | 25% (age $\geq 70$);<br>100% (age $< 70$) | 100% |
| 6. Combination | 100% | 25% (age $\geq 70$);<br>50% (age $< 70$) | 0% | 25% (age $\geq 70$);<br>50% (age $< 70$) | 65% |

**Table A2.** Probability distributions for sampling state transition times in the ABM (RM).

| Maturing out from state ... | ... into state | Probability distribution |
| --- | --- | --- |
| Susceptible | Exposed | N/A |
| Exposed | $I_p$ | $\sim \text{Gamma}(\text{mean } \mu=4.0, \text{ shape } k=4.0)$ |
| Exposed | $I_s$ | $\sim \text{Gamma}(\mu=4.0, k=4.0)$ |
| $I_p$ | $I_c$ | $\sim \text{Gamma}(\mu=1.5, k=4.0)$ |
| $I_s$ | Recovered | $\sim \text{Gamma}(\mu=5.0, k=4.0)$ |
| $I_c$ | Recovered | $\sim \text{Gamma}(\mu=3.5, k=3.5)$ |
| $I_c$ | H-ICU | $\sim \text{Gamma}(\mu=7.0, k=7.0)$ |
| $I_c$ | H-Non-ICU | $\sim \text{Gamma}(\mu=7.0, k=7.0)$ |
| H-ICU | Recovered | $\sim \text{Gamma}(\mu=10.0, k=10.0)$ |
| H-ICU | Dead | $\sim \text{Gamma}(\mu=10.0, k=10.0)$ |
| H-Non-ICU | Recovered | $\sim \text{Gamma}(\mu=8.0, k=8.0)$ |
| H-Non-ICU | Dead | $\sim \text{Gamma}(\mu=8.0, k=8.0)$ |

**Table A3.**
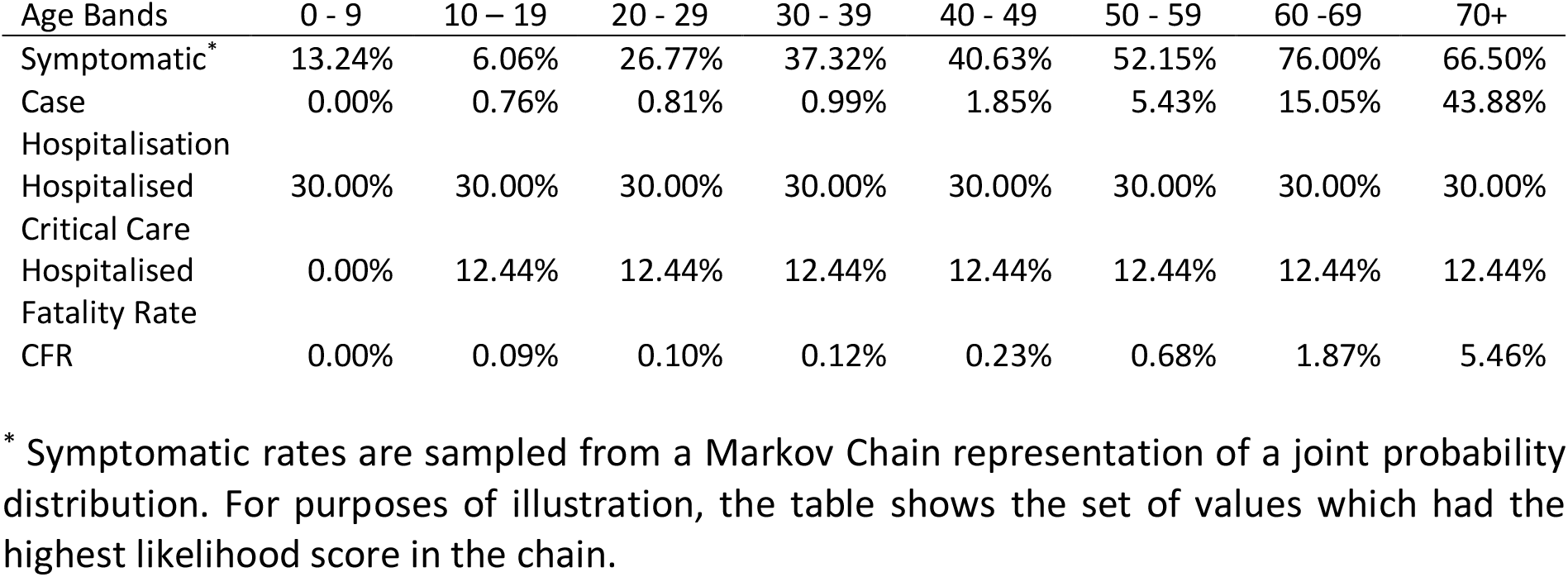
Age-based rates for routing between states in the ABM (RM), together with the Adjusted Case Fatality Rate (CFR) from Davies *et al*. (2020, Table S2).

## Footnotes

1 Continuity of the modelling team, for example due to reliance on researchers on temporary contracts or fractional contributions from grants is a general problem of large-scale modelling. The issue of flawed simulations has also long been recognised in the public debate about the hazards of economic modelling (Mason 2016, P. W. 2014).

2 The code and data files for the TM were downloaded from https://github.com/cmmid/covid-UK on 27 May 2020. The code scripts for the experiments are in R, while the core simulation engine is in C++.

3 This creates a conceptual problem as the contact rates for a composite category cannot mean the same as they do for specific categories. Someone on a bus and someone in a supermarket both fall into the “other” category but clearly do not, in fact, have contact.

4 Neither do the contact matrices account for the extent to which contact partners are repeated. In our experience, contacts made at home are largely with the same family members each day. Similarly, classmates and officemates vary little from day to day. Contact partners in the street or at the shops may, however, vary a great deal.

5 In addition, during school holiday periods, school contact is again assumed to be 0%.

6 Later experiments in Davies *et al*. (2020) explored the impact of delaying the launch of the intervention by 2, 4 or 8-week shifts.

7 All code and supporting files for the RM, together with data and scripts for the analyses in section 5, have been published at: https://github.com/innovative-simulator/CovidModelAgentBasedReproduction

8 *p* for IP, *s* for IS and *c* for IC to avoid nested subscripts that are practically illegible.

9 I3 signifies three distinct infectious states.

10 Note that, although requiring here 1+10=11 random numbers, our RM method is still more efficient than sampling for each agent whether that agent is being infected. That method would require 100 Bernoulli-distributed random numbers for this time step.

11 To follow Davies *et al*. (2020) in simulating every county in the UK, we would have needed to be able to simulate the largest county, West Midlands, which has 2.9 million agents. We tested our NetLogo model up to 500,000 agents with no problems other than computer time and the need to extend NetLogo’s JAVA heap size beyond its 1Gb default.

12 The number of points outside the percentiles is not large, but nonetheless we suspect this imperfection is real even though it may be less severe than it appears.

13 We communicated this bug to the authors and were told that, as it happened, they were already aware of it and that they planned to publish a correction in the same journal. For more details, see https://github.com/cmmid/covid-uk/issues/9

14 Again, the reproduction process focuses the mind on specific empirics. Counties are administrative units for data collection and do shape *some* forms of social contact (hospitals and state schools are almost always used on the basis of county of residence) but other social contacts (like family, friendship, shopping and recreation) are fairly unlikely to be shaped by them.

## Notes

### Competing Interest Statement

The authors have declared no competing interest.

### Funding Statement

No relevant funding.

### Author Declarations

Exemption. N/A to research content.

